# Preoperative optimisation in fast-track and enhanced recovery after surgery (ERAS) programs for total hip and knee joint replacement: a systematic review protocol

**DOI:** 10.1101/2021.09.27.21264189

**Authors:** Wei-Ju Chang, Justine Naylor, Victor Liu, Masiath Monuja, Sam Adie

## Abstract

**Introduction:** Emerging evidence suggests that fast-track and enhanced recovery after surgery (ERAS) targeting modifiable risk factors reduce complications after total hip (THR) and knee replacement (TKR). However, what constitutes an effective preoperative optimisation protocol for THR and TKR remains unclear. The aims are to: (1) describe pre-operative optimisation protocols for THR and TKR; (2) evaluate the effects of pre-operative optimisation protocols for THR and TKR on morbidity, and patient-reported outcomes.

**Methods and analysis:** Systematic review and meta-analysis. Electronic databases will be searched using pre-determined search terms to identify relevant studies and evaluate the study eligibility and risks of bias. Two independent reviewers will select the eligible studies and any disagreement will be resolved through a third reviewer. We will include studies investigated pre-operative optimisation protocols administered prior to participants receiving primary THR or TKR to improve post-operative outcomes. Primary outcomes are hospital readmission, complications and patient-reported pain and function. Risk of bias will be assessed using the Cochrane RoB 1 tool and strength of evidence will be examined using the GRADE approach. Pre-operative optimisation protocols will be summarised qualitatively. Meta-analyses on the effects of included protocols will be conducted if appropriate.

**Ethics and dissemination:** This systematic review does not require ethics approval. The findings will be disseminated in a peer-reviewed journal and presented at relevant conferences.

**Registration:** This protocol has been submitted to the International Prospective Register of Systematic Reviews on 30 August 2021.

**ARTICLE SUMMARY:** *Strength and Limitations:*

- This systematic review aims to describe pre-operative optimisation protocols for total hip and knee replacement and to synthesise evidence for the effects of pre-operative optimisation protocols on hospital readmission, complications and patient-reported outcomes.
- Two independent reviewers will conduct study selection, data extraction and risk of bias assessment.
- Meta-analyses, sub-group and sensitivity analyses will be performed where appropriate.

## INTRODUCTION

Osteoarthritis affects approximately 9.3% of the population in Australia, causing persistent pain, disability and reduced quality of life.^1^ The incidence of hip and knee osteoarthritis is increasing.^2 3^ While conservative (non-operative) management is successful for many people,^4^ total hip replacement (THR) and knee replacement (TKR) are recommended interventions for end-stage disease when non-operative interventions have failed.^5^

Although effective for improving pain, function and quality of life, THR and TKR are major surgeries that carry significant risks such as infection, deep vein thrombosis, implant failure, fracture that may require medical treatments or revision surgery.^6^ Research has identified several modifiable risk factors of post-operative complications, including obesity, diabetes, anaemia, smoking, use of opioids and vitamin D deficiency.^7-9^ In addition to adverse events, persistent post-operative pain and disability have been reported in subgroups of individuals after THR and TKR. Poor patient-reported outcomes are associated with some potentially modifiable physiological and psychological factors.^10-14^ Thus, some studies have investigated pre-operative interventions that aim to optimise THR/TKR outcomes via targeting known modifiable risk factors.^15-17^

The concept of pre-operative multimodal/multidisciplinary optimisation is not new, and the scope has expanded to incorporate principles of shared decision making.^18-23^ Although relatively sparse compared with other surgical specialities, optimisation programs for THR and TKR, such as pre-operative rehabilitation (prehabilitation), fast-track or enhanced recovery after surgery (ERAS) protocols have been developed and tested.^24-28^ A recent review of optimisation prior to knee replacement concluded that there is some, though limited, evidence that optimisation may improve outcomes post-surgery.^17^ Indeed, prehabilitation through patient education and exercise can improve patient-reported outcomes such as shorter hospital stay after THR/TKR, less postoperative pain and function after THR, greater quadriceps muscle strength after TKR.^16^ Further, emerging evidence suggests that pre-operative interventions targeting modifiable risk factors reduce the incidence of complications after THR and TKR.^29 30^ A recent systematic review has shown that ERAS protocols significantly reduce the length of hospital stay after THR and TKR, and treatment costs, although the effect on mortality rate remains unclear.^15^ However, pre-operative optimisation was not embedded in all included studies and the intervention details (i.e., type, duration, frequency of interventions) were not clearly revealed. Similarly, a recent consensus statement for perioperative care in THR and TKR provided recommendations for pre-operative optimisation but the details of the interventions were absent,^31^ and some recommendations (i.e., cessation of alcohol consumption and smoking) were not based on data from fully implemented ERAS for THR/TKR.^32^ It remains unclear what constitutes an effective pre-operative optimisation protocol for THR and TKR.

This systematic review aims to: (1) describe the pre-operative optimisation protocols for THR and TKR that are being used; (2) evaluate the effects of pre-operative optimisation protocols in isolation for THR and TKR on morbidity including hospital readmission and complications, and patient-reported outcomes.

## METHODS AND ANALYSIS

This systematic review protocol is prepared according to the Preferred Reporting Items for Systematic Reviews and Meta-Analysis Protocols (PRISMA-P) Guidelines.^33 34^ The PRISMA-P checklist is provided in the *Appendix (Table S1)*. Registration of this systematic has been submitted and is currently assessed by the International Prospective Register of Systematic Reviews (PROSPERO).

### Eligibility Criteria

#### Types of participants

Studies including adults aged 18 or older receiving primary elective THR or TKR surgeries will be included. The surgeries can be either unilateral or bilateral joint replacement. No restriction will be placed on sex or race. Studies including participants receiving partial joint replacements (i.e., unicompartmental or hemi-replacements) or joint replacements indicated for fracture, will not be included.

#### Types of intervention

Eligible studies will investigate pre-operative optimisation protocols administered *prior* to participants receiving a primary elective THR or TKR, with/without a comparative group that received a standard care protocol. No restriction will be placed on the duration of pre-operative optimisation protocol. Only studies had a comparative group will be used for achieving aim 2 of this review. The optimisation protocols should be intended to improve *post-operative* outcomes. To provide a comprehensive review on pre-operative optimisation, studies will be included if the intervention group received a continuum care program involving pre-, peri- or post-operative optimisation protocols (i.e., fast-track protocols). Only studies compared a pre-operative optimisation protocol with standard care will be used to achieve aim 2.

#### Type of outcome measures

The primary outcomes of this review for assessing aim 2 are: 1) hospital readmissions within 90 days; and 2) any complication including surgical site or other infection, cardiovascular event, venous thromboembolism, or mortality. The secondary outcomes include length of hospital stay, patient-reported pain and function outcomes. No restriction will be placed on when the outcomes were measured in the studies.

#### Types of studies

For Aim 1 of this review, we will include prospective randomised controlled trials (RCTs), prospective non-randomised clinical trials, prospective observational studies, and retrospective studies. For Aim 2, only prospective RCTs or prospective non-randomised clinical trials will be included in meta-analyses. Systematic or literature reviews, case reports or series, or conference abstracts will be excluded.

### Search strategy

To identify eligible, published studies, we will search the following electronic databases:

- MEDLINE
- EMBASE
- Cumulative Index to Nursing and Allied Health Literature (CINAHL)
- Cochrane Central Register of Controlled Trials (CENTRAL)

Search strategies will be established using medical subject headings (MeSH) and related text words and tailored to each database. A combination of different keywords for THR, TKR and pre-operative optimisation protocols will be used to identify relevant studies. The full search terms and search strategy are included in the *Appendix*. No restriction will be placed on the publication period, but only studies in the English language will be included. We will search the reference lists of eligible studies and relevant reviews to include any missed but relevant published studies. Citation searching for forward citation of recent studies and citation alerts (i.e., Google Scholar) on included studies will also be used to identify new studies as they appear during the review progress. We will re-run the searches prior to the final analysis to retrieve and include any relevant studies.

### Study selection

The EndNote X9 software (Clarivate Analytics) will be used to store, organise, and manage all search results and to remove duplicate records. Two reviewers will independently evaluate the title and abstract of all studies identified through the search against the eligibility criteria. The full text of the selected studies will then be retrieved. For studies retrieved from trial registrations, the full text is defined as all associated files and information. Any uncertainty about the study eligibility, the full text will be obtained for further information. Any disagreement over study eligibility will be resolved by consensus, and an additional reviewer will be consulted if needed. Excluded studies and the reasons for exclusion will be recorded and reported.

### Data extraction

A customised data extraction spreadsheet will be developed and piloted on two studies relevant to this review, before being used to extract data from the included studies. Two reviewers will independently perform data extraction from the final list of selected studies. Any disagreements in the extracted data will be resolved through discussion with a third reviewer. We will extract the following information from the included studies:

- *Study characteristics*: the first author, year of publication, study design, country, and study setting.
- *Participants*: age, sex, type of surgery (i.e., THR or TKR, unilateral or bilateral, left or right), duration of knee/hip osteoarthritis, co-morbidities, use of opioids, and the number of participants allocated in each intervention group.
- *Interventions*: details of the pre-operative optimisation protocol (i.e., type, duration, number of interventions, frequency, intervention providers), details of pre-, peri- or post-operative care protocols as a part of a continuum optimisation program if available.
- *Outcome measures*: the type of measure used to assess primary and secondary outcomes, at any peri- or post-operative timepoints.
- *Results*: data on the primary and secondary outcomes measured at any timepoints.

If data are missing, authors of the studies will be contacted a maximum of three times, after which the data will be considered irretrievable.

### Studies quality and risk of bias

Study quality and risk of bias will be assessed by two independent reviewers using the Cochrane Risk of Bias version 1 (RoB 1) tool for RCTs^35^ and the Cochrane Risk of Bias in Non-Randomised Studies - of Interventions (ROBINS-I) tool for non-randomised studies.^36^ Disagreement between reviewers will be resolved through discussion and a third reviewer will be consulted if consensus is not achieved. The risk of bias will be evaluated on the following domains: random sequence generation, allocation concealment, blinding of participants and personnel, blinding of outcome assessment, incomplete outcome data, selective reporting, and other potential sources of bias. The ROBINS-I tool will be used to assess the risk of bias for studies that have not used randomisation for intervention allocations (i.e., cross-sectional or cohort study designs). The ROBINS-I tool covers seven domains: confounding and participants’ selection (pre-intervention), intervention classification (during intervention), and deviations, missing data, measurements and selection of reported results (post-intervention).^36^ Both RoB 1 and ROBINS-I use signalling questions to guide judgments for each domain and an overall risk of bias assessment.

### Data synthesis

A narrative summary will be conducted to provide an overview of pre-operative optimisation protocols for THR and TKR respectively. The details of pre-operative optimisation protocols from all selected studies will be summarised in tables. The risk factors targeted by pre-operative optimisation protocols and the corresponding intervention provided will be listed.

For meta-analysis, only outcome data extracted from RCTs will be used and quantitatively synthesised using R (R version 4.0.0) software.^37^ A meta-analysis will be performed when there are at least two RCTs with a sufficiently similar population and intervention(s), measuring similar outcomes.^38^ Dichotomous outcomes (i.e., dichotomous complication or readmission outcomes) will be analysed as odd ratios (ORs) with 95% confidence intervals (CIs). Time-to-event outcomes will be analysed as hazard ratios (HRs) with 95% CIs. ORs and HRs will be calculated on a log scale and the results will be converted back to the original metric. When a trial reports different follow-up times that are summarised into the same category, the latest one will be used. Continuous outcomes (i.e., length of hospital stay, patient self-reported outcomes) will be analysed as mean differences with 95% CIs or standardised mean differences (SMD). The SMD for the combined effect estimate will be re-expressed on the scale of the original outcome measure used by the included studies.^39^ Heterogeneity will be assessed using the I^2^ statistic and the P value from the Chi^2^ test, and between study heterogeneity τ^2^.^40^ The P value < 0.1 and I^2^ > 50% indicate substantial heterogeneity. When substantial heterogeneity is present, a subgroup analysis will be performed to explore the potential sources of heterogeneity.

A narrative approach will be used to summarise the results from non-RCTs and retrospective studies based on study quality and type of surgery (THR/TKR). Study findings including data reduction, display and comparison, conclusion and the classification of evidence from individual studies will be summarised in a table.^41 42^

### Subgroup and sensitivity analysis

Where heterogeneity is identified (p < 0.1), subgroup analysis will be performed based on the type of surgery (THR vs. TKR). Further, a sensitivity analysis will be conducted to assess the impact of excluding studies with high risk of bias on the results of meta-analyses, if possible.

### Meta-biases

If more than 10 studies are included in meta-analysis, publication biases will be assessed using a funnel plot,^43^ and Egger’s test (p value > 0.10 indicating low publication bias).^44^ The risk of publication bias will be examined using a funnel plot asymmetry test, with a p value > 0.10 indicating low publication bias.^44^ To estimate the number of missing studies and to generate an adjusted estimate by imputing suspected missing studies, a trim and fill method, a non-parametric data augmentation approach, will be used.^45^ The adjusted estimates show whether the estimates based on meta-analysis are biased resulted from funnel plot asymmetry. A positive value of difference between unadjusted and adjusted estimates suggests that the estimate in meta-analysis is considered overestimated due to missing studies.^46^

### Quality of evidence

The Grading of Recommendations Assessment, Development, and Evaluation (GRADE) approach will be applied to rate the certainty of evidence and the strength of recommendations into four levels: very low, low, moderate, and high.^47^ The level of certainty of evidence will be downgraded for the following reasons:

- *Risk of bias*: The rating will be downgraded by two levels if there is a high “risk of bias” for > 25% and < 50% of the included studies participants. It will be one grade down, if > 50% of participants are from high risk of bias studies.^48^
- *Imprecision*: The rating will be downgraded by one level if the total number of participants included in meta-analyses is < 400 for continuous outcomes and < 300 for dichotomous outcomes.^49^
- *Inconsistency*: The rating will be downgraded by one level if significant heterogeneity is detected (p < 0.1).^50^
- *Indirectness*: This domain will not be considered as the eligibility criteria ensure specific populations and outcome interest.^51^
- *Publication bias*: The rating will be downgraded by one level if a publication bias is found via visual and statistical assessments.^52^

### Patient and public involvement

No patient involved.

## Data Availability

Not applicable.

## ETHICS AND DISSEMINATION

This review does not require ethics approval. The findings of the systematic review will be disseminated in a peer-reviewed journal and presented at scientific seminars and conferences in relevant fields such as orthopaedic surgery, rehabilitation or allied health.

## AUTHORS’ CONTRIBUTIONS

WJC, JN and SA conceptualised the study and developed the methodology of the review. WJC drafted the manuscript. All the authors reviewed and approved the final version of the manuscript.

## FUNDING STATEMENT

This research received no specific grant from any funding agency in the public, commercial or not-for-profit sectors.

## COMPETING INTEREST STATEMENT

The authors declare that they have no competing interests.

## APPENDIX

**TABLE S1.** PRISMA-P 2015 Checklist

| Section/topic | # | Checklist item | Information reported |  | Page number(s) |
| --- | --- | --- | --- | --- | --- |
|  |  |  | Yes | No |  |
| ADMINISTRATIVE INFORMATION |  |  |  |  |  |
| Title |  |  |  |  |  |
| Identification | 1a | Identify the report as a protocol of a systematic review | ☒ | ☐ | 1 |
| Update | 1b | If the protocol is for an update of a previous systematic review, identify as such | ☐ | ☐ | Not applicable |
| Registration | 2 | If registered, provide the name of the registry (e.g., PROSPERO) and registration number in the Abstract | ☒ | ☐ | 5, 6 |
| Authors |  |  |  |  |  |
| Contact | 3a | Provide name, institutional affiliation, and e-mail address of all protocol authors; provide physical mailing address of corresponding author | ☒ | ☐ | 1 |
| Contributions | 3b | Describe contributions of protocol authors and identify the guarantor of the review | ☒ | ☐ | 12 |
| Amendments | 4 | If the protocol represents an amendment of a previously completed or published protocol, identify as such and list changes; otherwise, state plan for documenting important protocol amendments | ☐ | ☐ | Not applicable |
| Support |  |  |  |  |  |
| Sources | 5a | Indicate sources of financial or other support for the review | ☒ | ☐ | 13 |
| Sponsor | 5b | Provide name for the review funder and/or sponsor | ☐ | ☐ | Not applicable |
| Role of sponsor/funder | 5c | Describe roles of funder(s), sponsor(s), and/or institution(s), if any, in developing the protocol | ☐ | ☐ | Not applicable |
| INTRODUCTION |  |  |  |  |  |
| Rationale | 6 | Describe the rationale for the review in the context of what is already known | ☒ | ☐ | 4, 5 |
|  |  |  | Yes | No |  |
| **Objectives** | 7 | Provide an explicit statement of the question(s) the review will address with reference to participants, interventions, comparators, and outcomes (PICO) | ☒ | ☐ | 5 |
| **METHODS** |  |  |  |  |  |
| **Eligibility criteria** | 8 | Specify the study characteristics (e.g., PICO, study design, setting, time frame) and report characteristics (e.g., years considered, language, publication status) to be used as criteria for eligibility for the review | ☒ | ☐ | 6 |
| **Information sources** | 9 | Describe all intended information sources (e.g., electronic databases, contact with study authors, trial registers, or other grey literature sources) with planned dates of coverage | ☒ | ☐ | 7 |
| **Search strategy** | 10 | Present draft of search strategy to be used for at least one electronic database, including planned limits, such that it could be repeated | ☒ | ☐ | 7, 8 |
| **STUDY RECORDS** |  |  |  |  |  |
| Data management | 11a | Describe the mechanism(s) that will be used to manage records and data throughout the review | ☒ | ☐ | 8 |
| Selection process | 11b | State the process that will be used for selecting studies (e.g., two independent reviewers) through each phase of the review (i.e., screening, eligibility, and inclusion in meta-analysis) | ☒ | ☐ | 8 |
| Data collection process | 11c | Describe planned method of extracting data from reports (e.g., piloting forms, done independently, in duplicate), any processes for obtaining and confirming data from investigators | ☒ | ☐ | 8, 9 |
| **Data items** | 12 | List and define all variables for which data will be sought (e.g., PICO items, funding sources), any pre-planned data assumptions and simplifications | ☒ | ☐ | 8, 9 |
| **Outcomes and prioritization** | 13 | List and define all outcomes for which data will be sought, including prioritization of main and additional outcomes, with rationale | ☒ | ☐ | 7 |
| **Risk of bias in individual studies** | 14 | Describe anticipated methods for assessing risk of bias of individual studies, including whether this will be done at the outcome or study level, or both; state how this information will be used in data synthesis | ☒ | ☐ | 9, 10 |
| **DATA** |  |  |  |  |  |
| **Synthesis** | 15a | Describe criteria under which study data will be quantitatively synthesized | ☒ | ☐ | 10 |
|  |  |  | Yes | No |  |
| | 15b | If data are appropriate for quantitative synthesis, describe planned summary measures, methods of handling data, and methods of combining data from studies, including any planned exploration of consistency (e.g., $I^2$, Kendall's tau) | ☒ | ☐ | 10 |
|  | 15c | Describe any proposed additional analyses (e.g., sensitivity or subgroup analyses, meta-regression) | ☒ | ☐ | 11 |
|  | 15d | If quantitative synthesis is not appropriate, describe the type of summary planned | ☒ | ☐ | 11 |
| **Meta-bias(es)** | 16 | Specify any planned assessment of meta-bias(es) (e.g., publication bias across studies, selective reporting within studies) | ☒ | ☐ | 11 |
| **Confidence in cumulative evidence** | 17 | Describe how the strength of the body of evidence will be assessed (e.g., GRADE) | ☒ | ☐ | 11, 12 |

### Search Strategies for Electronic Databases

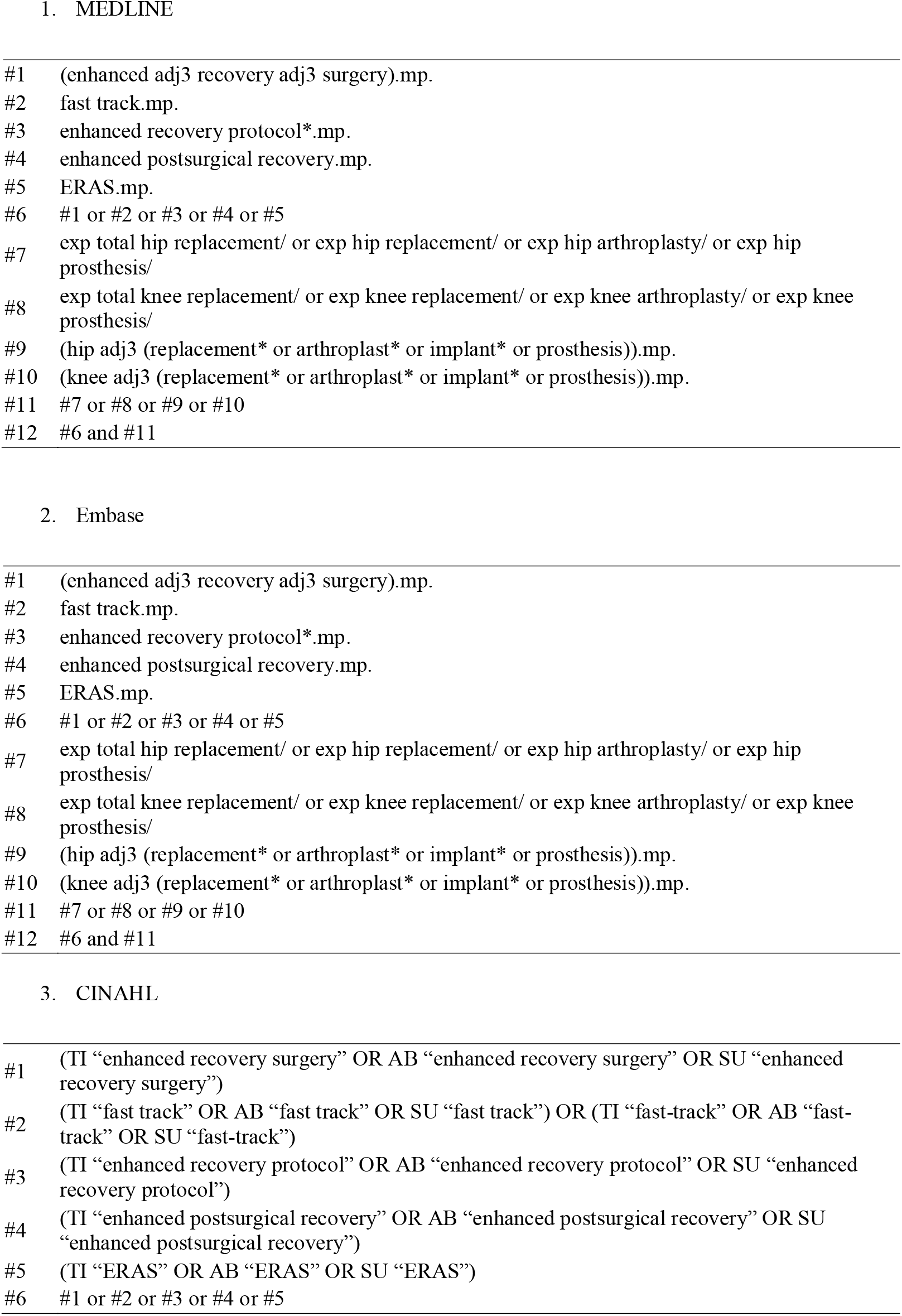
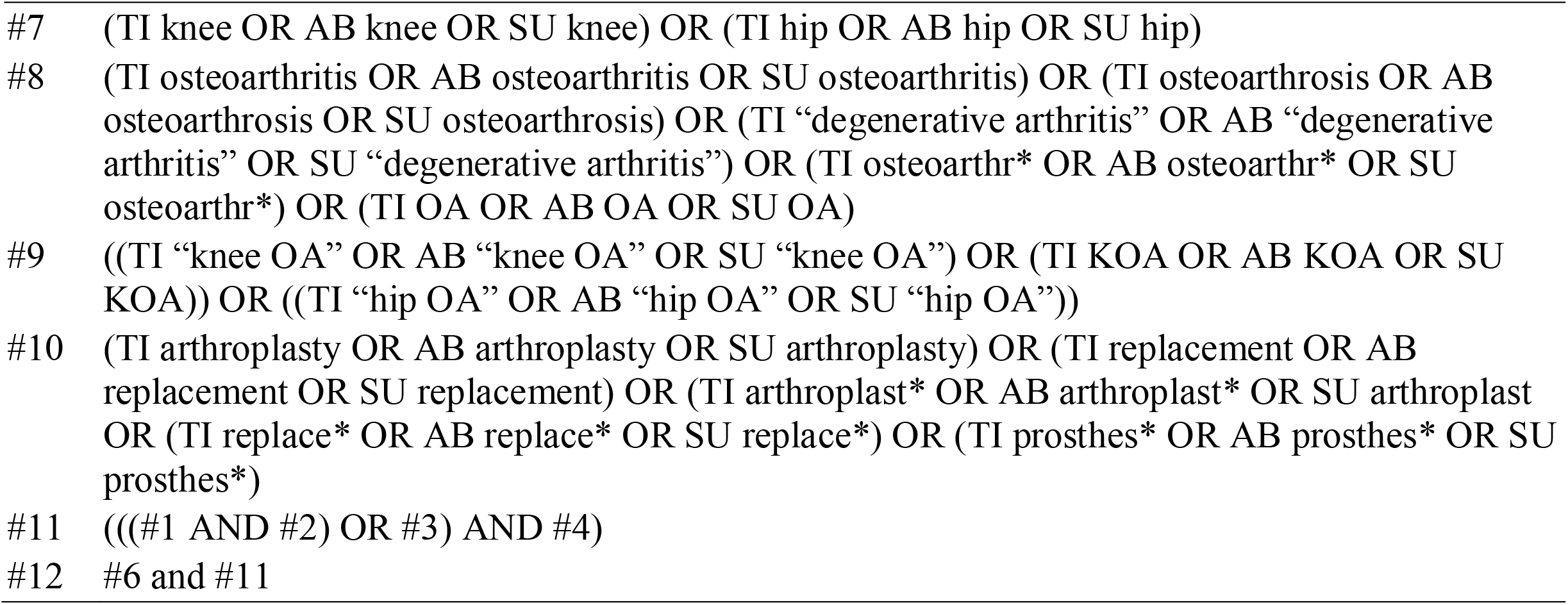

## Notes

### Competing Interest Statement

The authors have declared no competing interest.

### Funding Statement

No external funding was received.

### Author Declarations

This systematic review does not require an ethics approval.

